# Alternative splicing of *OAS1* alters the risk for severe COVID-19

**DOI:** 10.1101/2021.03.20.21254005

**Authors:** Jennifer Huffman, Guillaume Butler-Laporte, Atlas Khan, Theodore G. Drivas, Gina M. Peloso, Tomoko Nakanishi, Anurag Verma, Krzysztof Kiryluk, J. Brent Richards, Hugo Zeberg

**Affiliations:** Massachusetts Veterans Epidemiology Research and Information Center (MAVERIC), VA Boston Healthcare System, Bostoon, MA, USA, 02130; Departments of Medicine, Human Genetics, Epidemiology, Biostatistics and Occupational Health, McGill University, Lady Davis Institute, Jewish General Hospital, Montréal, Québec, Canada; Division of Nephrology, Department of Medicine, Vagelos College of Physicians & Surgeons, Columbia University, New York, NY; Department of Genetics, Perelman School of Medicine, University of Pennsylvania, Philadelphia, PA 19104, USA; Division of Human Genetics, Department of Pediatrics, Children’s Hospital of Philadelphia, Philadelphia; Department of Biostatistics, Boston University School of Public Health, Boston, MA 02118; Institute for Molecular Medicine Finland, Univerisity of Helsinki, Helsinki, Finland; Department of Human Genetics, McGill University, Montréal, Québec, Canada; Lady Davis Institute, Jewish General Hospital, McGill University, Montréal, Québec, Canada; Kyoto-McGill International Collaborative School in Genomic Medicine, Graduate School of Medicine, Kyoto University, Kyoto, Japan; Research Fellow, Japan Society for the Promotion of Science, Tokyo, Japan; Institute for Genomic Medicine, Columbia University, New York, NY; Department of Twin Research, King’s College London, London, UK; Max Planck Institute for Evolutionary Anthropology, Deutscher Platz 6, D-04103 Leipzig, Germany; Department of Neuroscience, Karolinska Institutet, SE-17177 Stockholm, Sweden

## Abstract

A locus containing *OAS1/2/3* has been identified as a risk locus for severe COVID-19 among Europeans ancestry individuals, with a protective haplotype of ∼75 kilobases derived from Neanderthals. Here, we show that among several potentially causal variants at this locus, a splice variant of *OAS1* occurs in people of African ancestry independently of the Neanderthal haplotype and confers protection against COVID-19 of a magnitude similar to that seen in individuals without African ancestry.

## MAIN TEXT

The COVID-19 pandemic has haunted the world for over a year. During this period, several large international efforts^1–4^ have been launched to identify the genetic determinants of COVID-19 susceptibility and severity. These efforts have identified more than a dozen genomic regions associated with severe COVID-19. However, the causal variants in these regions are yet to be identified, hampering our ability to understand COVID-19 pathophysiology.

When risk haplotypes are long it is more challenging to disentangle causal variants due to linkage disequilibrium (LD). This is especially problematic for haplotypes derived from Neanderthals and Denisovans that often span several tens of kilobases or more. Two notable COVID-19 examples are the major risk locus on chromosome 3 (3p21.31) and the *OAS1/2/3* locus on chromosome 12 (12q24.13), which both carry haplotypes of Neanderthal origin^5,6^. The *OAS* genes encode enzymes catalyzing the synthesis of short polyadenylates, which activate ribonuclease L that in turn degrades intracellular double-stranded RNA and triggers several other antiviral mechanisms^7^. The protective Neanderthal-derived haplotype confers ∼23% reduced risk of becoming critically ill upon infection with SARS-CoV-2^3^. Supporting this, a recent Mendelian randomization study found that increased circulating levels of OAS1 were associated with reduced risk of very severe COVID-19, hospitalization for COVID-19 and susceptibility to this disease^8^. However, other evidence from a transcription-wide association study, suggested a stronger association with OAS3 levels^3^. Thus, efforts are required to disentangle the causal gene, or genes, at this locus.

The *OAS* region was identified as a COVID-19 risk locus in association studies^1,3^ of mainly Europeans. The protective haplotype is derived from Neanderthals, is ∼75 kilobases long and covers the three genes *OAS1/2/3*^6^. A candidate causal variant in the region is rs10774671, which falls in a splice acceptor site of *OAS1* and where the protective (G) allele results in a longer and more active OAS1 enzyme^9^. However, this variant is as associated with COVID-19 severity as any of the hundreds of variants in LD. In European ancestry individuals we find 130 variants co-segregating (r^2^>0.8) with the splice-acceptor variant (**Fig. 1a**). Thus, further methods are required to disentangle the causal SNP(s) at this locus. Doing so, could help to identify the causal gene.

**Fig 1.**
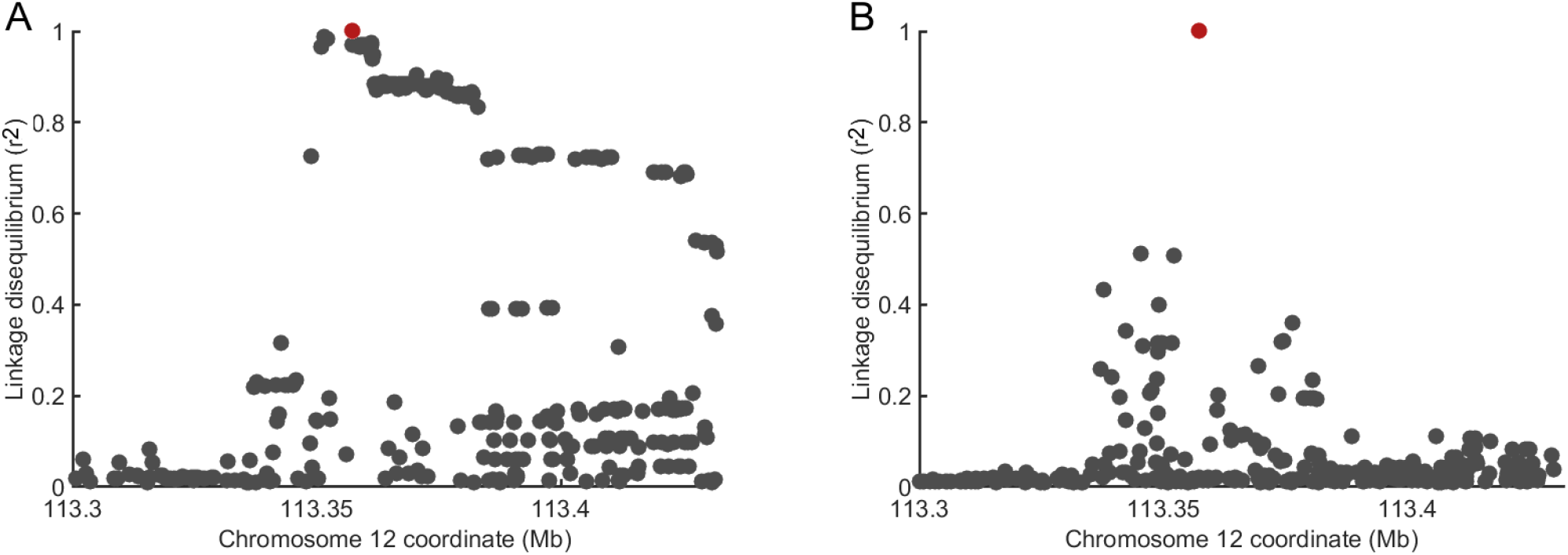
Linkage disequilibrium of the splice acceptor variant in individuals of European and African ancestries. **A**) Linkage decay in individuals European ancestry (n = 503). 130 variants are in LD (r^2^ > 0.8) with the splice acceptor variant rs10774671 (marked in red). **B**) Same as in A) but for African ancestry (n = 661). No variants were found to be in LD with the splice acceptor variant. Data from the 1000 Genomes project^10^. *X*-axis gives *hg19* coordinates.

One method to better identify causal SNPs at an associated locus is to test associations in different ancestries, particularly when these other populations have different LD structure and shorter haplotypes. Therefore, to examine if we could identify a population with which we could test this variant independently, we investigated the presence of co-segregating variants in populations in the 1000 Genomes project^10^. In South Asians, there are 129 such variants and in East Asian 128 variants. In stark contrast, no variants co-segregate with rs10774671 in Africans at a LD of r^2^>0.6 (**Fig. 1b**). Thus, the African ancestry population offers a possibility to independently test if rs10774671 is associated with COVID-19 severity.

To test the association of splice acceptor variant rs10774671 in people of African ancestry with COVID-19 outcomes we combined five studies that had assessed COVID-19 severity (UK BioBank, Penn Medicine BioBank, Columbia University COVID-19 Biobank, Biobanque Québec COVID-19 and the VA Million Veteran Program), comprising 1,842 cases and 118,631 controls of African Ancestry. We found that the rs10774671-G allele conferred a protection against COVID-19 hospitalization in this population (**Fig. 2**, p = 0.03) of similar magnitude (OR = 0.92, 95% confidence interval [CI]: 0.86-0.99) as in Europeans (OR = 0.89, 95% CI: 0.86-0.93), a population in which the rs10774671-G allele is less common (35% allele frequency versus 66% among African ancestry individuals^10^). Moreover, we find no evidence of heterogeneity accross the five studies (Cochran’s Q = 2.00, p = 0.74; I^2^ = 0.0% [0.0%-58.4%]; τ^2^ = 0.00 [0.00-0.09], 95% CI in brackets, see Methods). Thus, rs10774671 is associated with COVID-19 severity independently of the variants with which it is associated in non-African populations.

**Fig 2.**
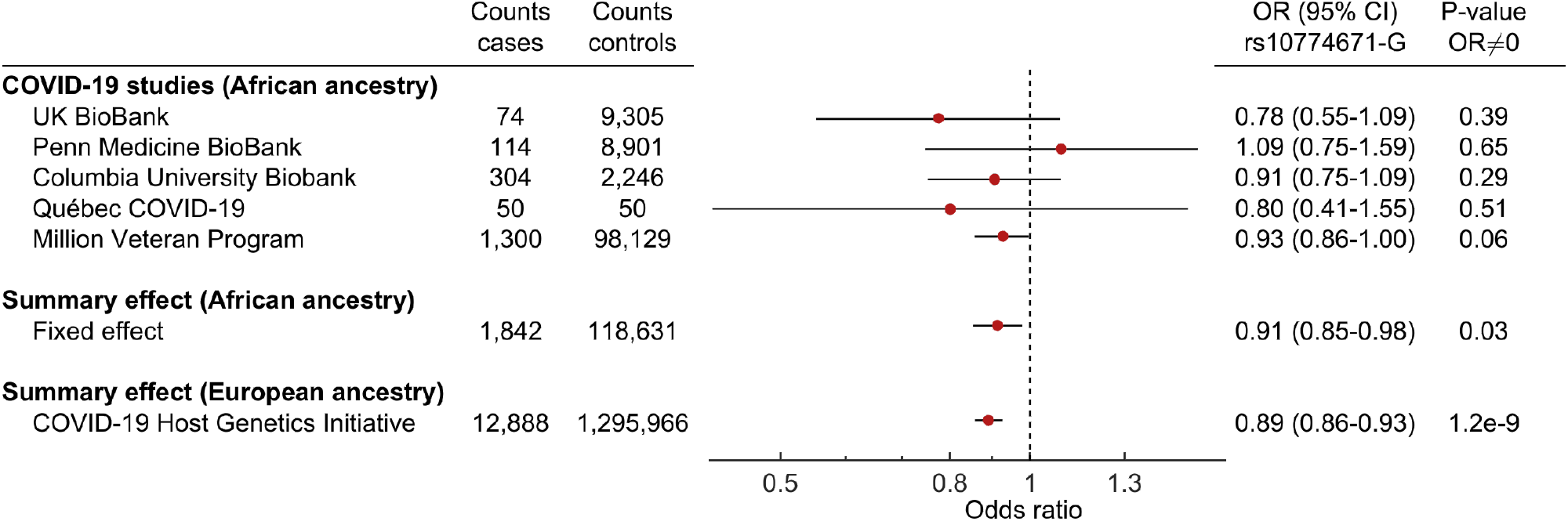
Odds ratios for COVID-19 hospitalization for African ancestry carriers of the ancestral splice variant. Summary effect in African ancestry individuals by a fixed effect meta-analysis of the five cohorts. Error bars give 95% confidence intervals.

This observation is compatible with the fact that Neanderthal haplotypes are rare or absent in African populations^11,12^ and that ancestral alleles seen in Neanderthals, such as the G allele at rs10774671, can also exist today as a result of inheritance from the ancestral population common to both modern humans and Neanderthals. In the latter case, such variants have existed in modern humans in the order of half a million years ago and therefore co-segregate with different variants than when they are derived from gene flow from Neanderthals into modern humans that occurred about 60,000 years ago^13^. Here, we leverage this fact to show that the ancestral splice variant, encoding a longer and more active enzyme^9^, is responsible for the protective effect associated with this locus^14^. These findings provide evidence that the splice-site variant at this locus influences COVID-19 outcomes by manipulating splicing of *OAS1*. Further, this rapid insight highlights the importance of including populations of different ancestries in genetic association studies and rapidly sharing data through large, international consortia.

## Data Availability

All data necessary to reproduce the findings can be found within the manuscript.

https://www.covid19hg.org/

https://www.internationalgenome.org/

## ACKNOWLEDGMENTS

We thank the COVID-19 Host Genetics Initiative and Regeneron for making the data from the genome-wide association study available. Genotyping and phenotyping of the Columbia University cohort was made possible by the Columbia University COVID-19 Biobank Genomics Workgroup members, including Richard Mayeux, Muredach P. Reilly, Wendy Chung, David B. Goldstein, Christine K. Garcia, Iuliana Ionita-Laza, Andrea Califano, Sheila M. O’Byrne, Danielle Pendrick, Soumitra Sengupta, Peter Sims, and Anne-Catrin Uhlemann. The Columbia University COVID-19 Biobank was supported by Columbia University and the National Center for Advancing Translational Sciences, NIH, through Grant Number UL1TR001873. The content is solely the responsibility of the authors and does not necessarily represent the official views of the NIH. The Richards research group is supported by the Canadian Institutes of Health Research (CIHR) (365825 and 409511), the Lady Davis Institute of the Jewish General Hospital, the Canadian Foundation for Innovation (CFI), the NIH Foundation, Cancer Research UK, Genome Québec, the Public Health Agency of Canada, the McGill Interdisciplinary Initiative in Infection and Immunity and the Fonds de Recherche Québec Santé (FRQS). GBL is supported by a CIHR scholarship and a joint FRQS and Québec Ministry of Health and Social Services scholarship. TN is supported by a research fellowship of the Japan Society for the Promotion of Science for Young Scientists. JBR is supported by an FRQS Clinical Research Scholarship. Support from Calcul Québec and Compute Canada is acknowledged. TwinsUK is funded by the Welcome Trust, the Medical Research Council, the European Union, the National Institute for Health Research-funded BioResource and the Clinical Research Facility and Biomedical Research Centre based at Guy’s and St. Thomas’ NHS Foundation Trust in partnership with King’s College London. The Biobanque Québec COVID19 is funded by FRQS, Genome Québec and the Public Health Agency of Canada, the McGill Interdisciplinary Initiative in Infection and Immunity and the Fonds de Recherche Québec Santé. HZ is supported by Jeanssons Stiftelser and Magnus Bergsvalls Stiftelse. We thank Svante Pääbo for careful reading of the manuscript and helpful comments. These funding agencies had no role in the design, implementation or interpretation of this study. The Penn Medicine BioBank (PMBB) is funded by a gift from the Smilow family, the National Center for Advancing Translational Sciences of the National Institutes of Health under CTSA Award Number UL1TR001878, and the Perelman School of Medicine at the University of Pennsylvania. We would like to acknowledge PMBB team members who made this work possible, including Daniel Rader, Marylyn Ritchie, Yuki Bradford, Shefali Setia Verma, Anastasia Lucas, Binglan Li.

## METHODS

### Study participants

Our analysis pooled hospitalized COVID-19 patients of African ancestry (n = 1,842) from five cohorts. The UK BioBank cohort contained 74 cases and 9,305 controls, the Penn Medicine BioBank 114 cases and 8,901 controls, the Columbia University Biobank 304 cases and 2,246 controls, Québec Covid-19 50 cases and 50 controls, and VA Million Veteran Program 1,300 cases and 98,129 controls. All participants gave appropriate consent and ethical approvals were obtained from the relevant research ethics boards.

### *Summary statistics* - VA *Million Veteran Program*

The VA Million Veteran Program (MVP) is a US-based longitudinal research program investigating how genes, lifestyle, and military exposures influences health and illness in Veterans, with study recruitment commencing in 2011^15^. Study participants were genotyped using a customized Affymetrix Axiom biobank array (the MVP 1.0 Genotyping Array), containing 723,305 variants^16^. Imputation was performed to a hybrid imputation panel comprised of the African Genome Resources panel (https://imputation.sanger.ac.uk/?about=1#referencepanels) and 1000G v3p5. COVID-19 cases were identified using an algorithm developed by the VA COVID National Surveillance Tool (NST)^17^. COVID-19-related hospitalizations were defined as hospital admissions between 7 days before and 30 days after an individual’s positive SARS-CoV-2 test. The association of hospitalized COVID-19 cases versus all other MVP participants was tested under an additive logistic model and was corrected for age, age^2^, sex, age-by-sex, and ethnicity-specific PCs. Individuals who died before March 1, 2020 were excluded as was one individual from each related pair. The analysis was restricted to only African American MVP participants (as defined by HARE^18^) resulting in 1,300 cases and 98,129 controls.

### Summary statistics - Biobanque Québécoise de la COVID-19

The Biobanque Québécoise de la COVID-19 (BQC-19) is a prospective hospital-based biobank recruiting patients with proven or suspected COVID-19 (Jewish General Hospital research ethics board no. 2020-2137). Whole genome genotyping was performed for all participants, with imputation using the TOPMed server. Individuals with African ancestry were determined by projecting genetic principal components on the 1000G reference panel. Our 50 cases were defined as patients hospitalized with COVID-19 or who died from the infection. Controls were the 50 other African ancestry participants, of which 32 had a clinical presentation consistent with COVID-19, but never had a positive test. An additive logistic regression model with the first 10 genetic principal components, age, sex, age^2^, age-by-sex, and age^2^-by-sex as covariates was used to determine the effect of the protective G allele on the risk of being a case.

### Summary statistics - Penn Medicine Biobank

The Penn Medicine Biobank (PMBB) contains ∼60,000 prospectively consented participants, all patients of Penn Medicine hospitals, for whom DNA samples have been obtained and on whom extensive phenotypic information has been generated from the electronic health record (EHR). 20,079 participants were genotyped using the Illumina Global Screening Array (Version 2) and further imputed using the TOPMed imputation server. SNPs with a call rate less than 1%, minor allele frequency less than 1%, or imputation info score less than 0.3 were excluded from further analysis. To define each ancestral group, principal component analysis was performed after merging the PMBB data with the 10000 Genomes Project reference dataset using the smartpca module of the Eigensoft package. We performed quantitative discriminant analysis (QDA) on all samples using the 1000 Genomes Project samples as a training sets to generate ancestry calls for all PMBB samples included in the analysis. Ultimately 9,015 African-ancestry genotyped samples were identified and included in our association study. All PMBB participants were followed for SARS-CoV-2 infection and hospitalization, with COVID-19 infection defined as any patient with a positive SARS-CoV-2 nasal swab or for whom the ICD billing code U07.1 had been coded in the EHR, and with COVID-19-related hospitalizations defined as the subset of these patients who had been admitted to hospital in the previous year with U07.1 as the admission diagnosis code, or who had been admitted for COVID-19-related symptoms as determined by manual chart review. Association analyses were performed using the Firth logistic regression test as implemented in REGENIE, including as covariates age, age^2^, sex, age-by-sex, and the first six ancestry-specific principal components of the genomic data. The PMBB is approved under Institutional Review Board of Perelman School of Medicine at University of Pennsylvania.

### Summary statistics - Columbia University Biobank

The Columbia University COVID-19 Biobank was established in response to the New York City infection surge in March 2020. The biobank recruited COVID-19 cases of diverse ancestry among all patients who were treated at Columbia University Irving Medical Center between March and May 2020. All cases were diagnosed by positive SARS-CoV-2 PCR test based on nasopharyngeal samples. The mean age of cases was 62.89 years, and the percentage of females was 43%. DNA of whole blood samples was extracted using standard procedures and genotyping was performed using the Illumina Global Diversity Array (GDA) chip. The controls were genotyped using the Illumina Multiethnic Global Ancestry (MEGA) chip. The analysis of intensity clusters and genotype calls were performed in Illumina Genome Studio software; all SNPs were called on forward DNA strand and standard quality control (QC) filters were applied, including per-SNP genotyping rate > 95%, per-individual genotyping rate > 90%, minor allele frequency (MAF) > 0.01, and Hardy–Weinberg equilibrium (HWE) test p-value > 10-08 in controls. The duplicates and cryptic relatedness in the given cohort were determined and excluded based on the estimated pairwise kinship coefficients > 0.0884. After QC, the dataset consisted of 6,757 individuals (1,029 cases and 5,728 controls) genotyped for 1,096,321 SNPs with overall genotyping rate of 99.9%. The imputation analysis was performed using TopMed imputation server. A total of 13,439,413 common markers imputed at high quality (R2 > 0.8 and MAF > 0.01) were used in downstream analyses. To define the African ancestry cluster, we used PCA against 1000 Genomes reference populations followed by k-means clustering on significant PCs of ancestry. The African ancestry cluster contained 332 cases positive for SARS-CoV-2 and 2,246 population controls. Of the 332 African ancestry cases, 304 had severe COVID-19 requiring hospitalization. Among the 304 cases included, 78 (26%) had respiratory failure requiring intubation and invasive ventilatory support, and 86 (28%) died due to COVID-19. We then tested the effect of rs10774671-G on the risk of hospitalization using SAIGE (Scalable and Accurate Implementation of GEneralized mixed model), after adjustment for sex and five principal components of ancestry. The collection of samples to the Columbia University COVID-19 Biobank was approved by the Institutional Review Board (IRB) of Columbia University (IRB protocol number AAAS7370), while the genetic analyses were approved under Columbia University IRB protocol number AAAS7948.

### Summary statistics - UK BioBank

Association analyses were performed using the Firth logistic regression test implemented in REGENIE, including as covariates age, age^2^, sex, age-by-sex, age^2^-by-sex, and ten ancestry-informative principle components. Association analyses were performed using the Firth logistic regression test implemented in REGENIE, including as covariates age, age2, sex, age-by-sex, age2-by-sex, and ten ancestry-informative principle components. The data was downloaded at https://rgc-covid19.regeneron.com/results [2021-02-02].

### Meta-analysis

The meta-analysis was done using inverse-variance weighting in the R-package meta. Heterogeneity was measured using Cochran’s Q, Higgin’s & Thompson’s I^2^, and τ^2^ using the DerSimonian-Laird estimator.

### Linkage disequilibrium

Linkage disequilibrium was calculated using LDlink^19^ 4.1 in the genomic region 113.30-113.45 Mb (*hg19*) using data from the 1000 Genomes Project^10^.

## Notes

### Competing Interest Statement

The authors have declared no competing interest.

### Author Declarations

The collection of samples to the Penn Medicine Biobank was approved by the Institutional Review Board of Perelman School of Medicine at University of Pennsylvania. The collection of samples to the Columbia University COVID-19 Biobank was approved by the Institutional Review Board (IRB) of Columbia University (IRB protocol number AAAS7370), while the genetic analyses were approved under Columbia University IRB protocol number AAAS7948. All participants gave appropriate consent and ethical approvals were obtained from the relevant research ethics boards.

### Summary of Updates

Author affiliations updated. Information regarding Penn Medicine Biobank updated.

